# Is Exercise During Androgen Deprivation Therapy Effective and Safe? A Randomized Controlled Trial

**DOI:** 10.1101/2025.05.05.25326963

**Authors:** Lauri Rantaniemi, Ilkka Jussila, Aino Siltari, Juha P. Ahtiainen, Annastiina Hakulinen, Eeva Harju, Jorma Sormunen, Tupu Nordström, Teuvo LJ Tammela, Teemu J Murtola

**Affiliations:** Faculty of Medicine and Health Technology, Tampere University, Tampere, Finland; Kanta-Häme Central Hospital; Department of Surgery, Wellbeing Services County of Central Finland, Jyväskylä, Finland; Faculty of Sport and Health Sciences, University of Jyväskylä, Jyväskylä, Finland; Department of Pharmacology, Faculty of Medicine, University of Helsinki, Helsinki, Finland; TAYS Cancer center, Tampere University Hospital, Tampere, Finland; Department of Gastroenterology, Department of Surgery, Tampere University Hospital, Wellbeing Services County of Pirkanmaa, Tampere, Finland; Docrates Cancer Center, Helsinki, Finland; Varala Sports Institute, Tampere, Finland

**Keywords:** Androgen deprivation therapy, Prostate cancer, Exercise, Resistance training, Quality of life

## Abstract

To explore the benefits and safety of supervised and unsupervised exercise among localized and metastatic prostate cancer patients (PCa) during long-term androgen deprivation therapy (ADT). A total of 44 PCa patients were enrolled in this randomized controlled trial (RCT). Participants were randomized in a 1:1 ratio into the supervised exercise sessions group or unsupervised home-based exercise group for three months. The primary outcomes assessed included quality of life (QoL), body composition, and metabolic markers, which were measured at baseline, after 3 months, and 6 months. Muscle strength was evaluated exclusively in the supervised exercise group. The main statistical models used were the Mann-Whitney U-test for between-group comparisons and the Wilcoxon rank-sum test for within-group changes. No adverse events were reported during the exercise period. There were no significant differences in QoL, body composition, or metabolic profiles between the intervention and control groups. The supervised exercise group demonstrated significant improvement in emotional functioning (Z = -2.102, p = 0.036), and all exercise performance metrics (p < 0.001), with the most pronounced gains observed in the leg press (Z = -4.17, p < 0.001). Furthermore, a significant association was identified between strength improvements and enhanced self-evaluated physical function (p < 0.001). Supervised exercise is safe for patients with localized and metastatic PCa undergoing ADT, and leads to significant improvements in emotional well-being, and muscle strength, which translates to better self-reported physical function. Findings underscore the need for RCTs with longer intervention and follow-up periods on supervised exercise, especially in metastatic PCa patients.

**Clinical trial registration number:** #NCT04050397

**Key points:** Supervised exercise was safe for patients with localized and metastatic prostate cancer who were undergoing long-term androgen deprivation therapy.

The supervised exercise among both localized and metastatic prostate cancer patients seems to result in improvements in diverse performance and strength-related metrics.

## 1 Introduction

Prostate cancer (PCa) is one of the most prevalent malignancies affecting men, with 1.5 million new cases in the year 2022 globally [^1^]. Androgen deprivation therapy (ADT) suppresses PCa progression and is commonly used in advanced PCa and as an adjuvant to radiation therapy for localized high-risk PCa [^2^]. However, low testosterone during ADT can cause adverse effects, such as a decrease in quality of life (QoL), functional capacity, loss of lean mass, and an increase in fat mass [^3, 4^].

Current guidelines recommend aerobic exercise 3 times a week, as well as resistance training 2 times a week to reduce many adverse effects of ADT [^5^]. Exercise impacts positively the reproductive system, fatigue, depression, metabolic function, and musculoskeletal system function [^6^]. Epidemiological studies have associated exercise with improved overall and PCa-specific survival [^7^]. In addition, Lopez P. et al. meta-analysis has shown that fat mass may be negatively associated with survival, and low levels of muscle mass could be associated with PCa progression [^8^]. From clinical studies there is strong evidence for exercise reducing several of the adverse effects of ADT such as loss of muscle mass and strength, fatigue and declining physical function. A moderate level of evidence has been shown for exercise-induced improvements in depression and anxiety, bone loss, and sexual dysfunction [^9^]. Additionally, exercise improves body composition [^10^] and aerobic capacity [^11^] in PCa patients undergoing ADT. Even low-intensity exercise can yield some of the benefits [^12^]. Preclinical studies have shown that exercise could reduce tumor growth [^13^], modulate metabolism [^14^], decrease hypoxia [^15, 16^], and activate immune cells [^13^] in rodent models. However, the evidence on whether the benefits of exercise translate into improvements in QoL [^17–19^] and functional capacity [^19, 20^] among men with PCa is mixed. Therefore, additional randomized clinical trials (RCTs) are needed to draw robust conclusions about the effectiveness of exercise in improving QoL and daily functioning.

The primary aim of this RCT is to assess the safety of both supervised and unsupervised exercise during ADT in PCa patients. Additionally, the study investigates the effects of exercise on QoL, daily activity levels, body composition, muscle strength, as well as serum glucose and lipoprotein levels.

## 2 Materials and Methods

### 2.1 Study Design and Participants

This randomized controlled pilot trial recruited 44 men undergoing long-term ADT for metastatic or non-metastatic PCa at Tampere University Hospital, Tampere, Finland. ADT was administered with GnRH antagonists or agonists, with or without the androgen receptor inhibitor bicalutamide and the androgen signaling inhibitors enzalutamide or darolutamide. Randomization was done by using a computer-generated random allocation sequence.

This study was registered at ClinicalTrials.gov under the identifier #NCT04050397 prior to the initiation of participant recruitment. Ethical approval for this trial was obtained from the Tampere University Hospital Ethics Board. Written informed consent was obtained from all participants before enrollment, emphasizing the voluntary nature of participation and the confidentiality of personal data.

Inclusion criteria were: 1) PCa patients on ADT during the study period; and 2) Signed informed consent. Exclusion criteria were: 1) Unable to participate in exercise (physical performance status ECOG 2 or higher); 2) High bone fracture risk, as judged by the clinician; and 3) Unable to understand spoken or written instructions in Finnish.

All study participants attended an introductory session, where a urologist explained the benefits of exercise during ADT, a licensed physical education instructor provided guidance on safe and effective home-based exercises, and a qualified dietician offered advice on proper food and nutrition. After the symposium, the men were randomized 1:1 to receive either 3 months of supervised exercise or home-based exercise (Figure I).

**Figure I.**
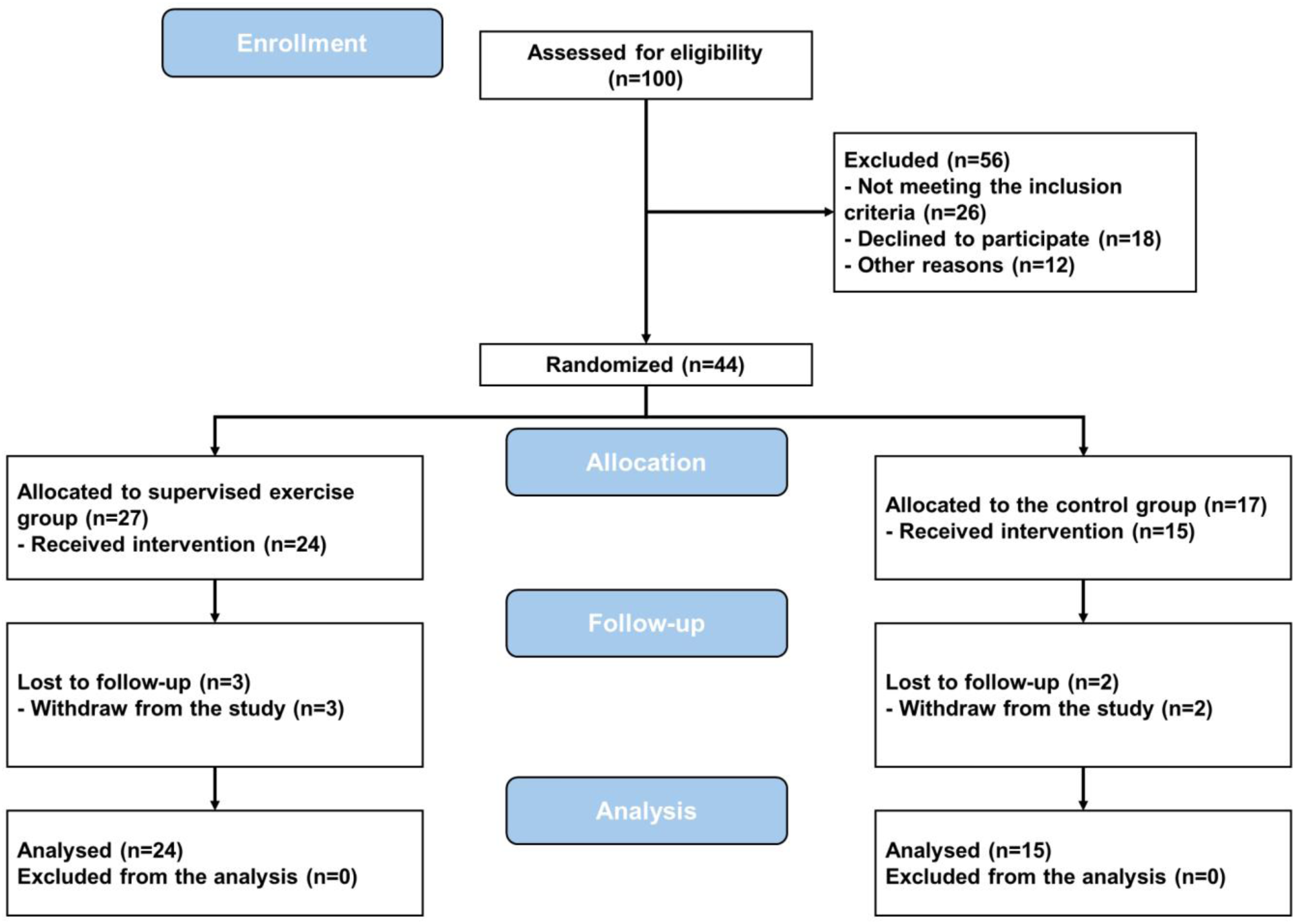
Study flow diagram.

### 2.2 Exercise Intervention

The exercise regimen for both groups during the study period is detailed in Table II. Participants in the intervention group attended a twice-weekly supervised exercise session at Varala Sports Institute, Tampere, Finland, and were also encouraged to complete a third weekly exercise session at home. Each supervised session consisted of 30 minutes of warm-up and 60 minutes of resistance training on exercise machines. The exercise regimen included seated rows, knee extensions, bench presses, core flexions, leg presses, and planks. The supervised exercise sessions followed the principles of progressive overload and were overseen by an exercise physiologist. In the first week, the number of repetitions per exercise was 15, followed by repetitions of 12 per exercise during the second week until the end of the intervention. The control group received an exercise instruction plan from a licensed exercise physiologist, encouraging them to follow it at home. The home-based exercise regimen was unsupervised, and adherence to exercises or time spent exercising were not monitored. After the three-month supervised exercise period, both groups were encouraged to continue exercising independently (Figure II).

**Figure II.**
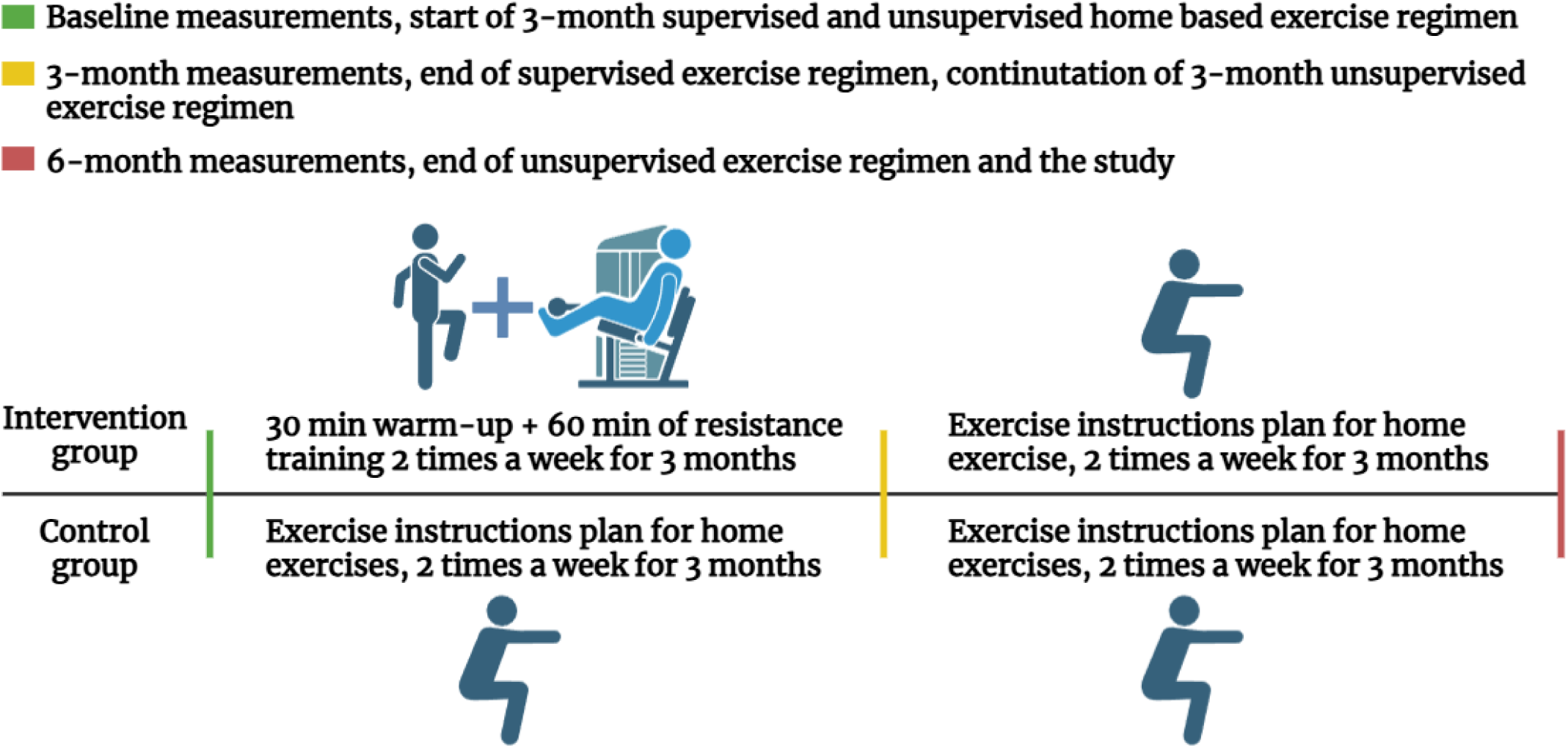
Exercise regimens for the intervention and control groups.

### 2.3 Data Collection

The data collection time points were at baseline, after 3 months, and 6 months of exercise (Table II). At all time points, the following data were collected: 1) QoL questionnaires: European Organization For Research And Treatment Of Cancer Core QoL questionnaire (ERTC QLQ-C30 and ERTC PR25); 2) Body composition, as measured with Tanita MC-980 body composition scale, including weight, body mass index (BMI), fat percentage and mass, visceral fat, and fat-free mass.; and 3) Blood samples, including fasting blood glucose, HbA1C, and lipoprotein profile (LDL, HDL, total cholesterol, and triglycerides) in the plasma. In addition, for the intervention group, seated row, knee extension, bench press, core flexion, leg press (measured in kilograms), and plank (measured in seconds) were recorded at baseline and after 3 months of supervised exercise.

### 2.4 Data analysis

QoL questionnaires were scored according to the EORTC scoring manual. ERTC QLQ-30 subdomain scores were divided into Global Health Status / QoL (QL), Physical Functioning (PF), Role Functioning (RF), Emotional Functioning (EF), Cognitive Functioning (CF), and Social Functioning (SF), Fatigue (FA), Nausea and Vomiting (NV), and Pain (PA), Dyspnoea (DY), Insomnia (SL), Appetite Loss (AP), Constipation (CO), Diarrhoea (DI), and Financial Difficulties (FI). The PR25 subdomain scores were divided into Sexual Activity (PRSAC) and Sexual Functioning (PRSFU), Urinary Symptoms (PRURI), Bowel Symptoms (PRBOW), Hormonal Treatment-Related Symptoms (PRHTR), and Incontinence Aid Use (PRAID). However, only the subdomain scores relevant to this trial were analyzed: ERTC QLQ-30: QoL, Physical Functioning (PF), Fatigue (FA), Nausea and Vomiting (NV), and Pain (PA). PR25: Sexual Activity (PRSAC) and Sexual Functioning (PRSFU), Urinary Symptoms (PRURI), Bowel Symptoms (PRBOW), Hormonal Treatment-Related Symptoms (PRHTR), and Incontinence Aid Use (PRAID).

All statistical analyses were performed using SPSS version 28. For the primary analysis, the absolute changes from baseline to 3 months and 0 to 6 months were calculated. Median changes were compared between the intervention arm and the control arm. The statistical model used for calculating U-, Z-, and p-values was the Mann-Whitney U-test. The Mann-Whitney U-test was the statistical model used for calculating U-, Z-, and p-values.

For the intervention group analysis, the median changes from baseline to 3 months and the delta change in percentage were calculated. The absolute values at baseline and 3 months were used in the statistical model. The statistical were performed using the Wilcoxon rank test, as well as Generalized Estimating Equations (GEE). In the intervention subgroup analysis, patients were stratified based on whether they experienced no strength gain (n = 10) or no increase in muscle strength (n = 14) during the intervention.

GEE was employed to evaluate only the effects of exercise and strength gain on physical and emotional function over time. Given the non-normally distributed nature of our data, we used the Hybrid method for parameter estimation, combining the advantages of Fisher scoring and Newton-Raphson methods to ensure robust and efficient convergence. The Pearson Chi-Square method was selected to estimate the scale parameter, which is appropriate for handling overdispersion in the data. Given the repeated measures design, we specified an unstructured working correlation matrix, allowing for different correlations at different time points.

## 3 Results

### 3.1 Patient characteristics

The baseline characteristics of participants in both the control and intervention groups are summarized in Table I. Prostate-specific antigen (PSA) levels were higher in the intervention group with a median PSA of 4.00 ng/mL, compared to 2.64 ng/mL in the control group (p-value here).

**Table I.**
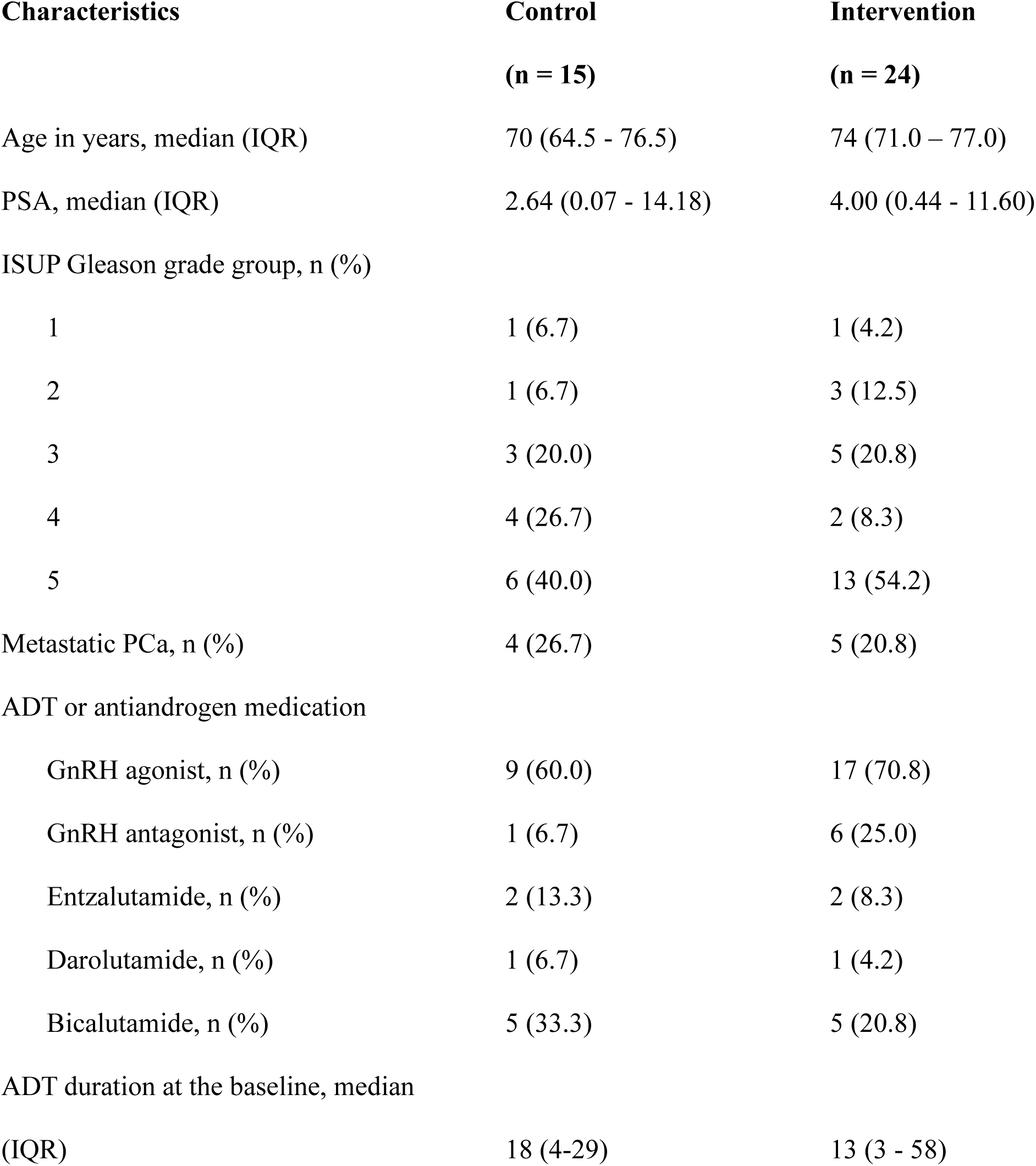
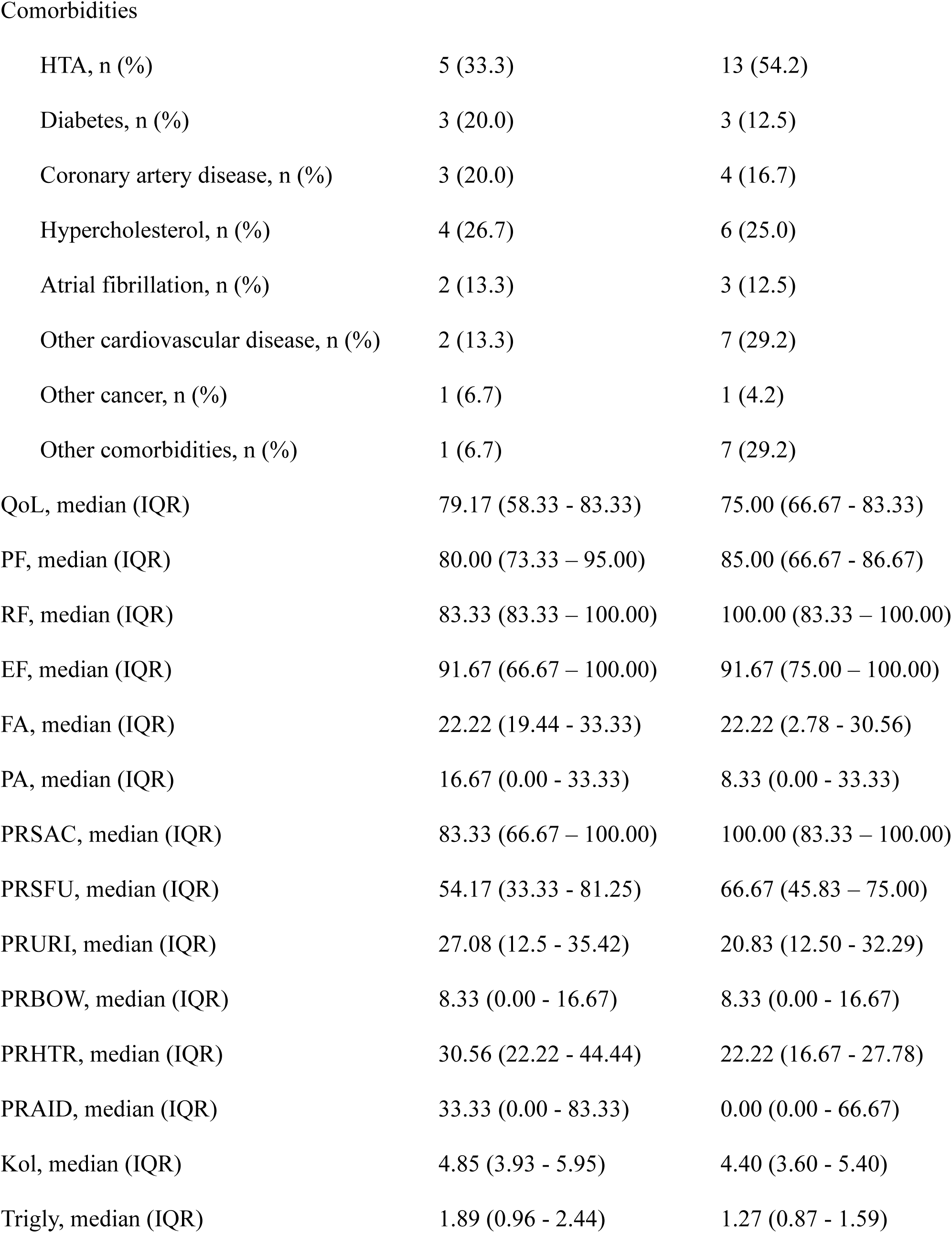
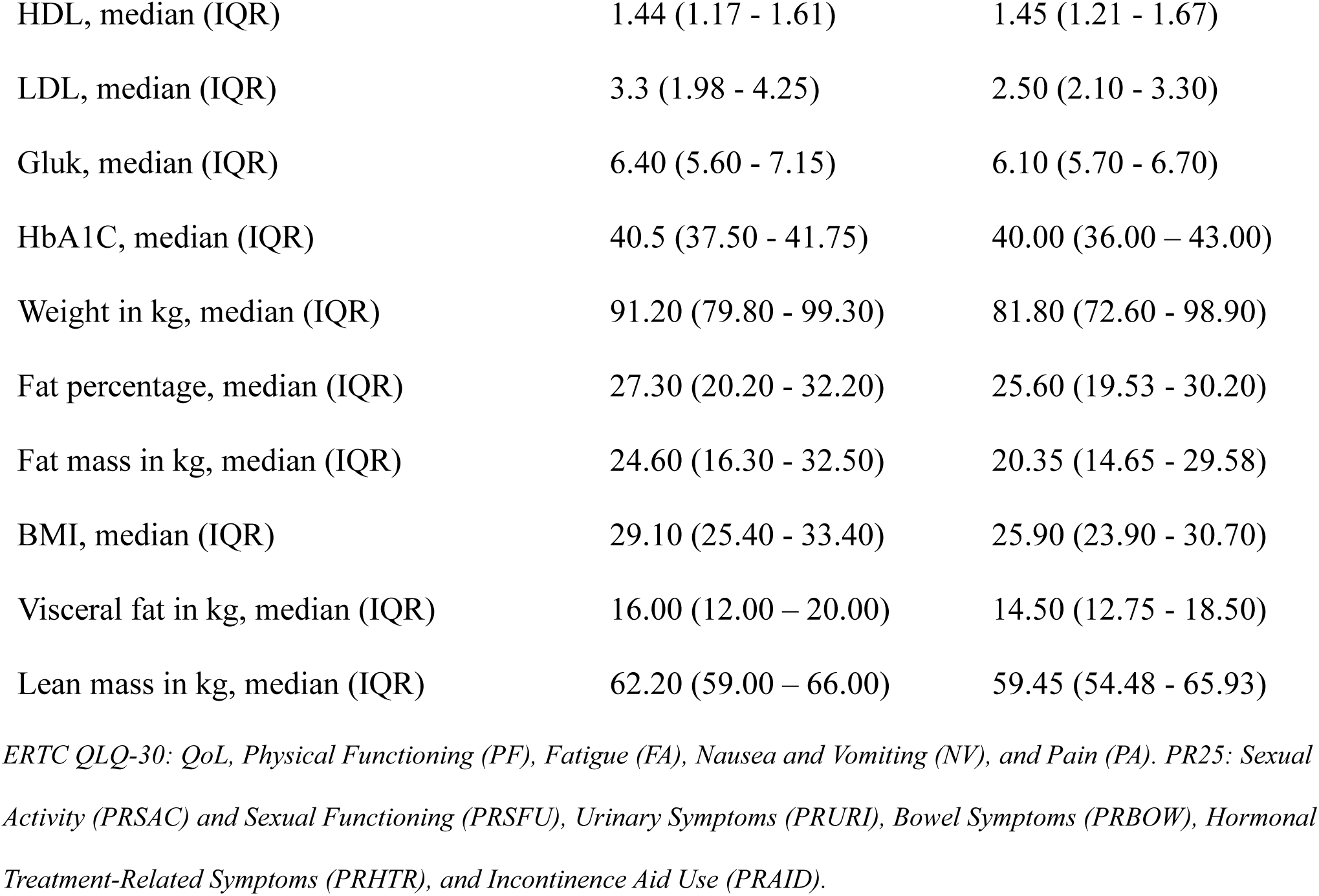
PCa patients baseline characteristics.

| Characteristics | Control<br>(n = 15) | Intervention<br>(n = 24) |
| --- | --- | --- |
| Age in years, median (IQR) | 70 (64.5 - 76.5) | 74 (71.0 – 77.0) |
| PSA, median (IQR) | 2.64 (0.07 - 14.18) | 4.00 (0.44 - 11.60) |
| ISUP Gleason grade group, n (%) |  |  |
| 1 | 1 (6.7) | 1 (4.2) |
| 2 | 1 (6.7) | 3 (12.5) |
| 3 | 3 (20.0) | 5 (20.8) |
| 4 | 4 (26.7) | 2 (8.3) |
| 5 | 6 (40.0) | 13 (54.2) |
| Metastatic PCa, n (%) | 4 (26.7) | 5 (20.8) |
| ADT or antiandrogen medication |  |  |
| GnRH agonist, n (%) | 9 (60.0) | 17 (70.8) |
| GnRH antagonist, n (%) | 1 (6.7) | 6 (25.0) |
| Entzalutamide, n (%) | 2 (13.3) | 2 (8.3) |
| Darolutamide, n (%) | 1 (6.7) | 1 (4.2) |
| Bicalutamide, n (%) | 5 (33.3) | 5 (20.8) |
| ADT duration at the baseline, median<br>(IQR) | 18 (4-29) | 13 (3 - 58) |
| Comorbidities |  |  |
| HTA, n (%) | 5 (33.3) | 13 (54.2) |
| Diabetes, n (%) | 3 (20.0) | 3 (12.5) |
| Coronary artery disease, n (%) | 3 (20.0) | 4 (16.7) |
| Hypercholesterol, n (%) | 4 (26.7) | 6 (25.0) |
| Atrial fibrillation, n (%) | 2 (13.3) | 3 (12.5) |
| Other cardiovascular disease, n (%) | 2 (13.3) | 7 (29.2) |
| Other cancer, n (%) | 1 (6.7) | 1 (4.2) |
| Other comorbidities, n (%) | 1 (6.7) | 7 (29.2) |
| QoL, median (IQR) | 79.17 (58.33 - 83.33) | 75.00 (66.67 - 83.33) |
| PF, median (IQR) | 80.00 (73.33 - 95.00) | 85.00 (66.67 - 86.67) |
| RF, median (IQR) | 83.33 (83.33 - 100.00) | 100.00 (83.33 - 100.00) |
| EF, median (IQR) | 91.67 (66.67 - 100.00) | 91.67 (75.00 - 100.00) |
| FA, median (IQR) | 22.22 (19.44 - 33.33) | 22.22 (2.78 - 30.56) |
| PA, median (IQR) | 16.67 (0.00 - 33.33) | 8.33 (0.00 - 33.33) |
| PRSAC, median (IQR) | 83.33 (66.67 - 100.00) | 100.00 (83.33 - 100.00) |
| PRSFU, median (IQR) | 54.17 (33.33 - 81.25) | 66.67 (45.83 - 75.00) |
| PRURI, median (IQR) | 27.08 (12.5 - 35.42) | 20.83 (12.50 - 32.29) |
| PRBOW, median (IQR) | 8.33 (0.00 - 16.67) | 8.33 (0.00 - 16.67) |
| PRHTR, median (IQR) | 30.56 (22.22 - 44.44) | 22.22 (16.67 - 27.78) |
| PRAID, median (IQR) | 33.33 (0.00 - 83.33) | 0.00 (0.00 - 66.67) |
| Kol, median (IQR) | 4.85 (3.93 - 5.95) | 4.40 (3.60 - 5.40) |
| Trigly, median (IQR) | 1.89 (0.96 - 2.44) | 1.27 (0.87 - 1.59) |
| HDL, median (IQR) | 1.44 (1.17 - 1.61) | 1.45 (1.21 - 1.67) |
| LDL, median (IQR) | 3.3 (1.98 - 4.25) | 2.50 (2.10 - 3.30) |
| Gluk, median (IQR) | 6.40 (5.60 - 7.15) | 6.10 (5.70 - 6.70) |
| HbA1C, median (IQR) | 40.5 (37.50 - 41.75) | 40.00 (36.00 – 43.00) |
| Weight in kg, median (IQR) | 91.20 (79.80 - 99.30) | 81.80 (72.60 - 98.90) |
| Fat percentage, median (IQR) | 27.30 (20.20 - 32.20) | 25.60 (19.53 - 30.20) |
| Fat mass in kg, median (IQR) | 24.60 (16.30 - 32.50) | 20.35 (14.65 - 29.58) |
| BMI, median (IQR) | 29.10 (25.40 - 33.40) | 25.90 (23.90 - 30.70) |
| Visceral fat in kg, median (IQR) | 16.00 (12.00 – 20.00) | 14.50 (12.75 - 18.50) |
| Lean mass in kg, median (IQR) | 62.20 (59.00 – 66.00) | 59.45 (54.48 - 65.93) |

*ERTC QLQ-30: QoL, Physical Functioning (PF), Fatigue (FA), Nausea and Vomiting (NV), and Pain (PA). PR25: Sexual* *Activity (PRSAC) and Sexual Functioning (PRSFU), Urinary Symptoms (PRURI), Bowel Symptoms (PRBOW), Hormonal* *Treatment-Related Symptoms (PRHTR), and Incontinence Aid Use (PRAID).*

Regarding disease aggressiveness, a higher percentage of participants in the intervention group had more aggressive disease (54.2% with ISUP Gleason grade 5) compared to the control group (40.0%). Both groups had a similar proportion of metastatic PCa (approximately 26%).

Regarding medications, GnRH agonists were more commonly used in the intervention group (70.8%) compared to the control group (60.0%), while GnRH antagonists were administered to 25.0% of the intervention group and only 6.7% of the control group. Enzalutamide use was similar between the groups, with 8.3% in the intervention group and 13.3% in the control group. Additionally, darolutamide was used by a small proportion of participants in both groups (4.2% in the intervention group and 6.7% in the control group). Bicalutamide, however, was more commonly used in the control group (33.3%) compared to the intervention group (20.8%).

Comorbid conditions such as hypertension were more prevalent in the intervention group (54.2% vs. 33.3%), while diabetes and coronary artery disease were relatively similar between the groups. QoL scores at baseline showed similar medians in both groups, with 79.17 (IQR: 58.33 - 83.33) for the control group and 75.00 (IQR: 66.67 - 83.33) for the intervention group. Physical function (PF) scores were also comparable, with medians of 80.00 (IQR: 73.33 - 95.00) in the control group and 85.00 (IQR: 66.67 - 86.67) in the intervention group.

The intervention group had lower lipoprotein level at baseline, compared to the control group (median LDL levels 2.50 mmol/L vs 3.3 mmol/L, triglycerides 1.27 mmol/L vs 1.89 mmol/L, cholesterol 4.40 mmol/L vs 4.85 mmol/L) as well as lower median weight (81.80 kg, IQR: 72.60 - 98.90) and BMI (25.90, IQR: 23.90 - 30.70) compared to the control group (91.20 kg, IQR: 79.80 - 99.30, and BMI of 29.10, IQR: 25.40 - 33.40, respectively).

### 3.2 Effect and safety of exercise

Table II presents the primary outcome, which were the changes from baseline after 3 and 6 months of exercise for QoL, lipoprotein profile, glucose, and body composition in both the control and intervention groups.

**Table II.**
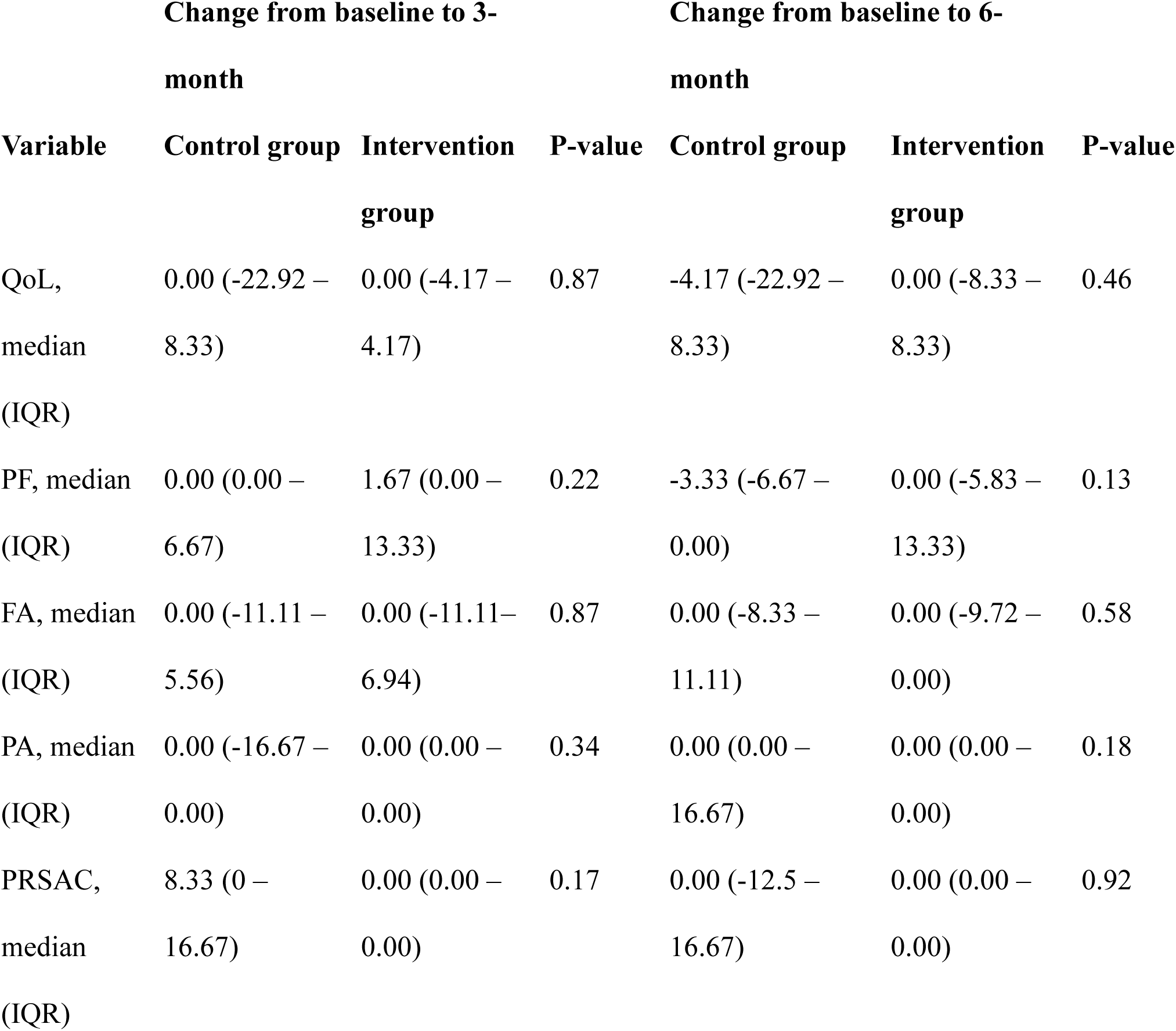
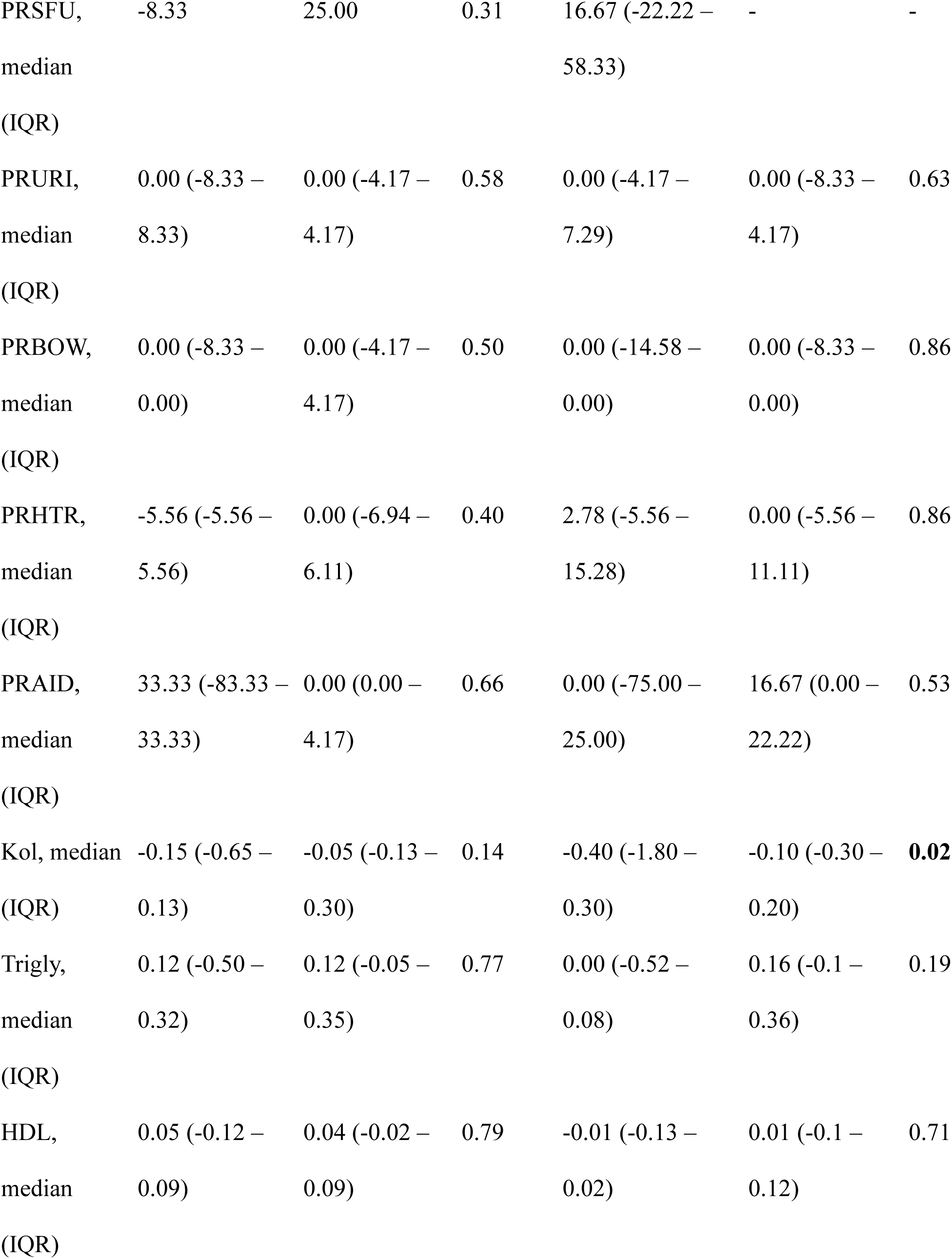
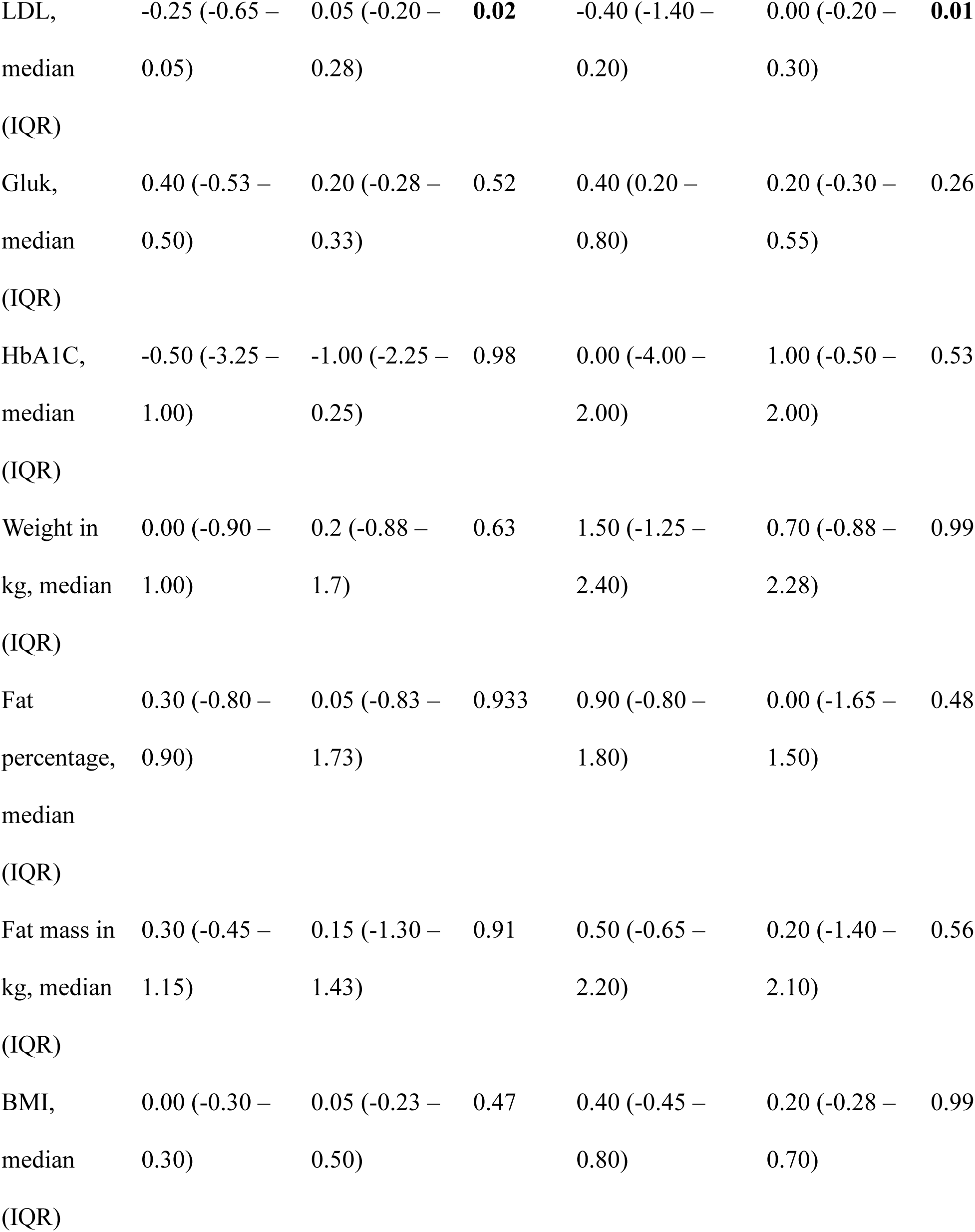
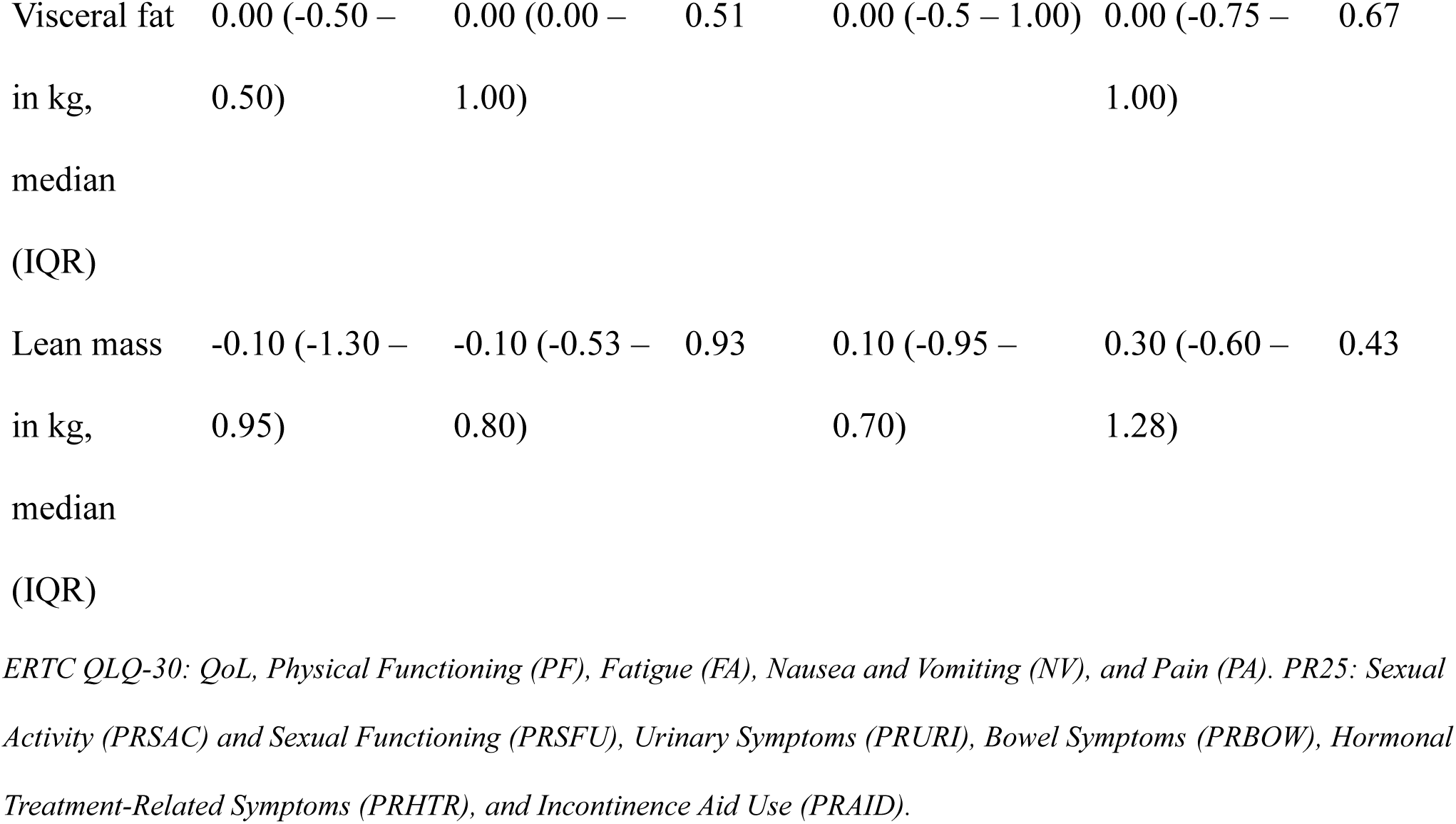
Intervention and control groups median change from baseline to 3- and 6-month.

A significant difference was found in LDL cholesterol levels between groups. At 3 months, the control group had a median change of -0.25 (IQR: -0.65 to -0.05) compared to 0.05 (IQR: -0.20 to 0.28) in the intervention group (U = 109.0, Z = 2.30, p = 0.022, r = 0.37). At 6 months, the control group had a median change of -0.40 (IQR: -1.40 to -0.20), while the intervention group experienced no change (IQR: -0.20 to 0.30) (U = 99.00, Z = 2.53, p = 0.012, r = 0.41). Total cholesterol levels also showed significant differences in the 6 months, with the control group experiencing a median change of -0.40 (IQR: -1.80 to -0.30) compared to -0.10 (IQR: -0.30 to 0.20) in the intervention group (U = 95.00, Z = 2.27, p = 0.024, r = 0.36). Exercise intervention did not change QoL, blood fasting glucose level, or body composition parameters significantly compared to controls.

In the secondary outcome, after 3 months of supervised training, the intervention group showed significant strength improvements across all measured resistance exercises (Figure III and Table III). For the seated row, a 98.65% increase was observed, and the Wilcoxon rank test yielded a Z-score of -4.165 (r = 0.85) with a p-value of less than 0.001, indicating a highly significant improvement. Similarly, significant improvements were found for knee extension (Z = -3.607, p < 0.001, r = 0.74, 61.34% increase), bench press (Z = -2.543, p = 0.011, r = 0.52, 31.30% increase), core flexion (Z = -3.709, p < 0.001, r = 0.76, 111.78% increase), leg press (Z = -4.173, p < 0.001, r = 0.85, 65.53% increase), and plank (Z = -3.753, p < 0.001, r = 0.77, 206.29% increase). These results demonstrate significant increases in the intervention group, with increases ranging from 31 % to 200% across all exercises (Supplementary Table I).

**Figure III.**
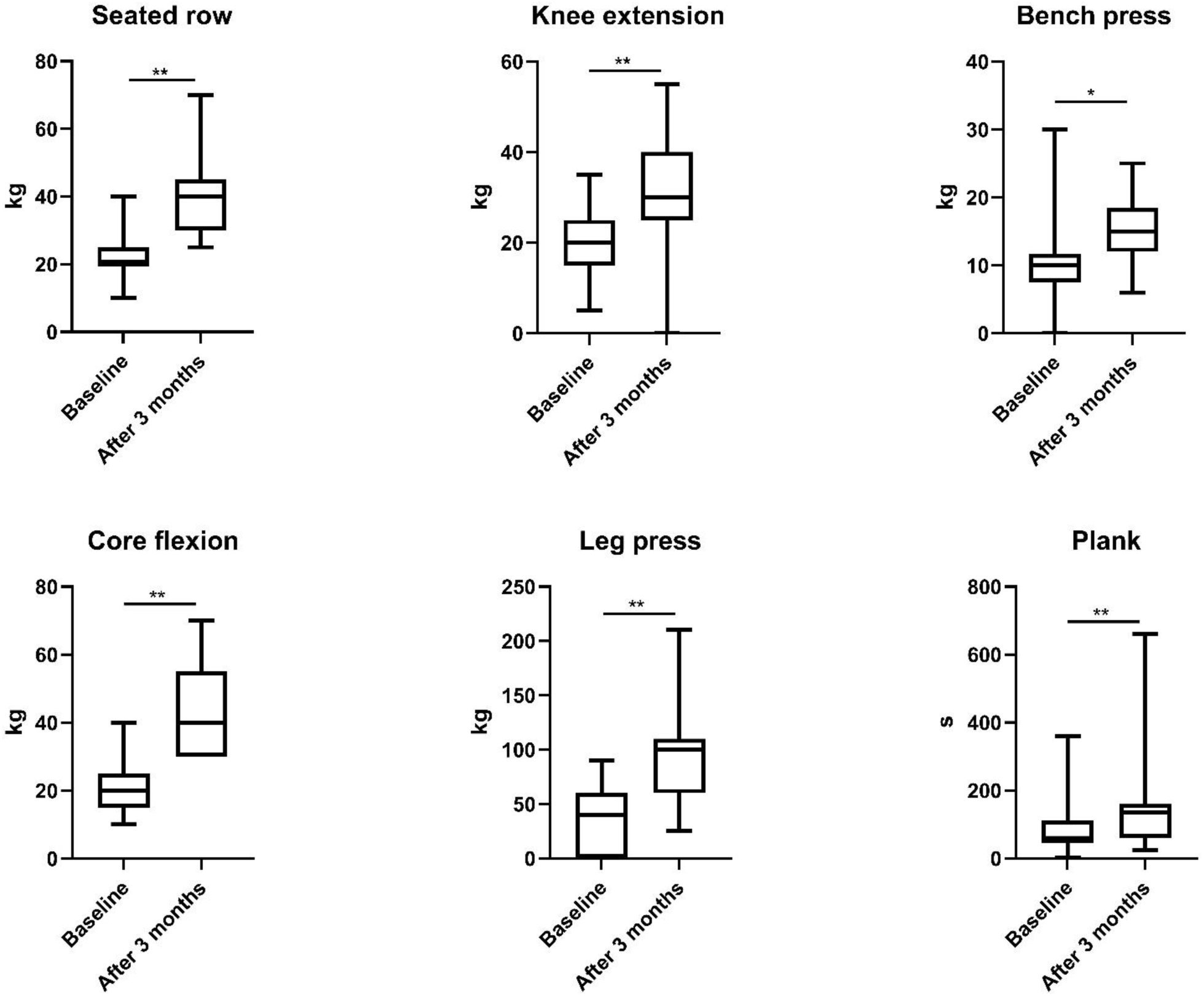
Box and whisker plot of the effects of supervised strength training on strength parameters (* = p = 0.05, ** = p < 0.001).

**Table III.** Wilcoxon rank test for intervention and intervention subgroups to determine the effectiveness of supervised strength training.

| Variable | Intervention group |  | No strength gain group |  | Strength gain group |  |
| --- | --- | --- | --- | --- | --- | --- |
|  | Z-score | P-value | Z-score | P-value | Z-score | P-value |
| Seated row | -4.165 | **<0.001** | - | - | - | - |
| Knee extension | -3.607 | **<0.001** | - | - | - | - |
| Bench press | -2.543 | **0.011** | - | - | - | - |
| Core flexion | -3.709 | **<0.001** | - | - | - | - |
| Leg press | -4.173 | **<0.001** | - | - | - | - |
| Plank | -3.753 | **<0.001** | - | - | - | - |
| GoL | -0.054 | 0.957 | -0.378 | 0.705 | -0.333 | 0.739 |
| PF | -1.580 | 0.114 | -0.170 | 0.865 | -2.226 | **0.026** |
| RF | -2.058 | **0.040** | -1.633 | 0.102 | -1.289 | 0.197 |
| EF | -2.102 | **0.036** | -0.137 | 0.891 | -2.461 | **0.014** |
| FA | -0.643 | 0.520 | -0.750 | 0.453 | -0.216 | 0.829 |
| PA | -0.087 | 0.931 | -0.447 | 0.655 | -0.276 | 0.783 |
| PRSAC | -0.351 | 0.726 | -0.816 | 0.414 | -0.378 | 0.705 |
| PRSFU | 0.000 | 1.00 | - | - | - | - |
| PRURI | -0.578 | 0.563 | -0.496 | 0.620 | 0.000 | 0.175 |
| PRBOW | -0.265 | 0.791 | -0.414 | 0.679 | -1.211 | 1 |
| PRHTR | 0.000 | 1.00 | -1.378 | 0.168 | -0.333 | 0.226 |
| PRAID | 0.000 | 1.00 | - | - | - | - |
| Cholesterol | -0.776 | 0.438 | -0.137 | 0.891 | -0.780 | 0.436 |
| Triglycerides | -1.413 | 0.158 | -0.631 | 0.528 | -1.332 | 0.183 |
| HDL | -1.351 | 0.177 | -1.992 | **0.046** | -0.631 | 0.528 |
| LDL | -0.805 | 0.421 | -0.412 | 0.680 | -0.736 | 0.462 |
| Glucose | -0.593 | 0.553 | -0.530 | 0.596 | -1.166 | 0.244 |
| HbA1C | -1.449 | 0.147 | -1.511 | 0.131 | -0.742 | 0.458 |
| Weight in kg | -1.026 | 0.305 | -0.280 | 0.779 | -1.644 | 0.100 |
| BMI | -1.009 | 0.313 | -0.426 | 0.670 | -1.693 | 0.090 |
| Whole body fat mass<br>in kg | -0.879 | 0.380 | -0.702 | 0.483 | -1.479 | 0.139 |
| Visceral fat mass in kg | -1.414 | 0.157 | 0.000 | 1 | -1.633 | 0.102 |
| Whole body lean mass<br>in kg | -0.224 | 0.823 | -0.170 | 0.865 | -0.384 | 0.701 |

Additionally, as a secondary outcome, emotional functioning improved in the intervention group significantly (Z = -2.102, p = 0.036, r = 0.43, 7.68% increase). However, role functioning showed a significant decline (Z = -2.058, p = 0.040, r = 0.42, -7.36% decrease) after 3 months of supervised exercise.

In secondary outcome a subgroup analysis of the intervention group, participants who gained strength in all exercises experienced a significant improvement in physical (Z = -2.226, p = 0.026, r = 0.60, 1.37% increase) and emotional function (Z = -2.461, p = 0.014, r = 0.66, 9.6% increase). In contrast, the subgroup with no strength gain showed no significant changes.

Table IV presents the results from the Generalized Estimating Equation (GEE) models estimating the effect of supervised strength training and strength gain on physical and emotional function. Exercise (time) did not have a significant effect on physical function (B = 0.05, SE = 0.04, p = 0.243). However, strength gain was a significant positive predictor of physical function (B = 0.22, SE = 0.06, p < 0.001), suggesting that patients who experienced strength gain had improved physical function over time.

**Table IV.** Generalized Estimating Equation Model estimating the effect of supervised strength training and strength gain on physical and emotional function.

| Dependent variable | Predictor | Estime (B) | SE | Wald Chi-Square | P-value |
| --- | --- | --- | --- | --- | --- |
| Physical function | Intercept | 4.22 | 0.08 | 3008.75 | 0 |
|  | Time | 0.05 | 0.04 | 1.58 | 0.243 |
|  | Strength gain | 0.22 | 0.06 | 12.28 | **<0.001** |
|  | Strength exercises |  |  |  |  |
|  | Intercept | 4.34 | 0.11 | 1649.60 | 0.000 |
|  | Time | -0.11 | 0.09 | 1.75 | 0.186 |
|  | Strength gain | 0.31 | 0.11 | 7.17 | **0.007** |
|  | Seated row | -0.02 | 0.01 | 8.07 | **0.005** |
|  | Knee extension | 0.01 | 0.01 | 3.65 | 0.056 |
|  | Bench press | -0.01 | 0.01 | 0.86 | 0.354 |
|  | Core flexion | 0.00 | 0.00 | 1.18 | 0.276 |
|  | Leg press | 0.00 | 0.00 | 1.85 | 0.173 |
|  | Plank | 0.00 | 0.00 | 3.89 | **0.049** |
| Emotional<br>function | Intercept | 4.38 | 0.07 | 4406.30 | 0.000 |
|  | Time | 0.06 | 0.03 | 3.30 | 0.069 |
|  | Strength gain | 0.03 | 0.05 | 0.25 | 0.621 |
|  | Strength exercises |  |  |  |  |
|  | Intercept | 4.38 | 0.06 | 5992.06 | 0.000 |
|  | Time | -0.01 | 0.06 | 0.01 | 0.909 |
|  | Strength gain | 0.01 | 0.04 | 0.02 | 0.903 |
|  | Seated row | 0.00 | 0.00 | 0.02 | 0.879 |
|  | Knee extension | 0.00 | 0.00 | 0.08 | 0.784 |
|  | Bench press | 0.01 | 0.00 | 3.76 | 0.052 |
|  | Core flexion | 0.00 | 0.00 | 0.58 | 0.448 |
|  | Leg press | 0.00 | 0.00 | 0.67 | 0.413 |
|  | Plank | 0.00 | 0.00 | 0.80 | 0.370 |

When specific resistance training exercises were included in the model, exercise remained a non-significant factor (B = -0.11, SE = 0.09, p = 0.186) while strength gain continued to be a significant positive predictor (B = 0.31, SE = 0.11, p = 0.007). Among the specific strength exercises, seated row had a small but significant negative effect on physical function (B = -0.02, SE = 0.01, p = 0.005), while plank showed a slight positive effect (B = 0.00, SE = 0.00, p = 0.049). Knee extensions, bench presses, core flexions, and leg presses did not have significant effects. However, the model did not identify any significant effect of exercise or strength gain on emotional function (p > 0.05).

## 4 Discussion

The main findings demonstrated that exercise was safe in PCa patients undergoing ADT, in both localized and metastatic PCa. Exercise did not significantly impact overall QoL, body composition, or glucose and lipid metabolism in either exercise group. Nonetheless, significant improvements were observed across all resistance training exercises in the supervised exercise group. Additionally, the supervised exercise group demonstrated significant improvements in emotional functioning. Further, strength gain was associated with an improvement in physical function. The adherence to activity monitoring was suboptimal, limiting the assessment of overall physical activity levels.

Previous systematic reviews and meta-analyses about the effects of exercise during ADT concluded that supervised exercise during ADT increases muscle strength [^21^], decreases fat mass, and increases lean mass [^22^] as well as may improve QoL and functional capacity of PCa patients [^23^]. However, our findings did not show any significant benefits of exercise on overall QoL. The sample size in this pilot trial was likely too small to detect significant differences in these parameters. Nonetheless, we demonstrated that the supervised exercise regimen was effective in improving strength, which translated into enhanced self-evaluated physical function even in a small trial such as ours. This suggests that strength gain has a robust influence on PCa patients’ QoL.

Improvement in strength and physical function is one of the most important benefits of exercise, which is crucial in maintaining functional independence [^24, 25^] and reducing bone fractures [^26^]. Additionally, we found that improvements in physical function occurred only in PCa patients who experienced strength gains, underscoring the importance of inter-individual variability in exercise adaptations among this population. This aligns with the backed-up previous findings, which have shown that healthy individuals experience inter-individual variance in resistance training adaptations [^27, 28^]. Inter-individual variability in resistance training adaptations has also been linked to differences in improvements in physical function [^29^], emphasizing the need to account for this variability when assessing PCa patients’ responses to exercise interventions.

The emotional functioning improvements may be attributed to the psychological benefits of regular exercise, such as reduced anxiety and depression [^30, 31^], which are common in PCa patients undergoing ADT [^32, 33^]. However, in our study, this effect is more likely attributable to the social aspect of the exercise, as this was observed only in the supervised exercise group, which participated in group exercise. This conclusion is strengthened by the finding that strength gain was not associated with the improvement in emotional function. Also, previous findings have shown that supervised group exercise is superior to home-based exercise in improving QoL [^34^]. One of the factors for superiority could be the social aspect of group exercise. The significant improvement in emotional functioning in the supervised exercise group highlights the need for a holistic approach to cancer care that includes not only physical but also psychological support. Group-based supervised exercise programs can provide social interaction and emotional support, which are beneficial for QoL [^31^].

The decline in role functioning suggests that while exercise may improve emotional well-being, it might not translate into perceived improvements in daily roles and activities, possibly due to the ongoing burden of cancer treatment and its associated symptoms. Previously, ADT has been shown to reduce the functional capacity of PCa patients [^35^].

The significant improvements in muscle strength observed in the intervention group underscore the importance of incorporating resistance training into cancer care for PCa patients undergoing ADT. Enhancing muscle strength is crucial for maintaining functional independence [^28, 29^], which can help prevent falls and fractures [^26^], a common risk for PCa patients [^36, 37^].

The finding that only patients who experienced strength gains also showed improvements in physical function highlights the necessity of personalized exercise regimens. Tailoring exercise programs to individual capabilities and monitoring progress closely can optimize the benefits of physical activity for PCa patients.

Our results show that even a relatively short, supervised exercise program led to meaningful improvements in emotional well-being and physical function, which suggests that clinicians could confidently recommend supervised exercise early during ADT to garner these benefits. We also emphasize the finding that supervised, group-based exercise had psychosocial benefits (improved emotional functioning) likely due to the social support element. This implies that clinicians should consider recommending group exercise opportunities or supervised sessions when possible. Furthermore, our findings reinforce current recommendations that PCa patients should engage in regular exercise during treatment to manage the adverse effects of treatment [^38^].

Our findings underscore the need for pragmatic strategies to support sustained exercise participation in men on long-term ADT, aligning with the PACC framework’s call for phase-specific approaches to exercise across the cancer care continuum [^39^]. In the future, larger trials with longer exercise interventions and extended follow-up periods should be conducted to see whether longer exercise periods yield superior results or maintain benefits achieved during the first couple of months, and to show how long the benefits last after the exercise period has ended.

### 3.3 Limitations

Several limitations of this study should be acknowledged. As the primary endpoint of this pilot trial was to evaluate the safety of exercise programs in PCa patients in ADT, the study did not include power or sample size calculations. This limitation means that the study may be underpowered to detect some differences in secondary endpoints, and the findings should be interpreted with caution. In the intervention group, only the number of repetitions was pre-determined, while the number of sets was not specified or monitored. This may have resulted in variations in the overall intensity and volume of exercise. However, the primary goal of the study was to pilot the feasibility of the exercise intervention in PCa patients. The home-based exercise regimen for the control group was unsupervised, potentially leading to differences in exercise adherence and intensity compared to the supervised intervention group. In addition, we did not monitor or record prior or current lifestyle habits such as diet or sleep, which could also affect exercise responses. Future research with predefined sample size calculations and adequate power is needed to draw robust conclusions about the effects of exercise in PCa patients, and to explore the synergistic benefits of medication and exercise as in the ongoing ESTRACISE [^40^] and MOVES (NCT05796973) trials.

## 5 Conclusion

In conclusion, this pilot RCT demonstrated that both supervised and home-based exercise is safe for localized and metastatic PCa patients undergoing ADT. Significant improvements in strength, physical function, and emotional functioning were observed exclusively in the supervised exercise group. These findings highlight the importance of integrating structured, supervised exercise programs into cancer care for PCa patients to enhance both physical and psychological well-being.

## 6 Perspective

Supervised exercise was safe for patients with localized and metastatic PCa undergoing ADT, and led to significant improvements in emotional well-being and muscle strength, which translated to better self-reported physical function. Findings underscore the need for larger randomized controlled trials with longer intervention and follow-up periods on supervised exercise, especially in metastatic PCa patients.

## Data Availability

Due to the high profile, and sensitive nature of the participants in this study, data cannot be shared according to the national laws in Finland.

## Abbreviations used in the article

ADT: Androgen Deprivation Therapy
PCa: Prostate Cancer
QoL: Quality of Life
EORTC QLQ-C30: European Organisation for Research and Treatment of Cancer Quality of Life Questionnaire
PR25: Prostate Cancer-Specific Quality of Life Module
ECOG: Eastern Cooperative Oncology Group Performance Status
GEE: Generalized Estimating Equations
IQR: Interquartile Range
PSA: Prostate-Specific Antigen
ISUP: International Society of Urological Pathology
GnRH: Gonadotropin-Releasing Hormone
HbA1C: Glycated Hemoglobin
LDL: Low-Density Lipoprotein
HDL: High-Density Lipoprotein
BMI: Body Mass Index
EORTC QLQ-30: Main Quality of Life Section of the Questionnaire
PRSFU: Sexual Functioning
PRSAC: Sexual Activity
PRURI: Urinary Symptoms
PRBOW: Bowel Symptoms
PRHTR: Hormonal Treatment-Related Symptoms
PRAID: Incontinence Aid Use

## 8 Ethics declarations

### 7.3 Ethics Approval

This study was registered at ClinicalTrials.gov under the identifier #NCT04050397 prior to the initiation of participant recruitment. Ethical approval for this trial was obtained from the Tampere University Hospital Ethics Board.

### 7.4 Consent to Participate

Written informed consent was obtained from all participants before enrollment, emphasizing the voluntary nature of participation and the confidentiality of personal data.

### 7.6 Author contributions

TJ Murtola, L Rantaniemi, A Siltari, A Hakulinen, E Harju, T Nordström conceptualized the research project, and design. L Rantaniemi, A Hakulinen, and T Nordström were involved in data collection for the study. I Jussila, L Rantaniemi, and A Siltari were responsible for the analysis. TJ Murtola, I Jussila, L Rantaniemi, A Siltari,, J Ahtiainen, J Sormunen, TLJ Tammela, and E Harju, were responsible for interpretation of the results. I Jussila, L Rantaniemi, J Sormunen drafted the manuscript. All authors critically reviewed and edited the manuscript prior to submission.

