## Supplementary Table I for "Is Exercise During Androgen Deprivation Therapy Effective and Safe? A Randomized Controlled Trial"

**Supplementary Table I**. Intervention group median baseline and post-supervised training values with Δ % change.

| Variable | Baseline | Post-training | Change (Δ %) |
|---|---|---|---|
| Seated row in kg, median (IQR) | 20 (20 – 25) | 40 (30 – 45) | 98.65 |
| Knee extension in kg, median (IQR) | 20 (15 – 25) | 30 (25 – 39) | 61.34 |
| Bench press in kg, median (IQR) | 10 (6 - 11) | 15 (12 – 19) | 31.30 |
| Core flexion in kg, median (IQR) | 20 (15 – 25) | 40 (34 – 56) | 111.78 |
| Leg press in kg, median (IQR) | 40 (0 – 60) | 100 (60 – 110) | 65.53 |
| Plank in seconds, median (IQR) | 60 (45 – 112) | 135 (60 – 160) | 206.29 |
| EORTC QLQ-C30 | | | |
| GL, median (IQR) | 75 (62.50 – 83.33) | 70.83 (66.66 – 83.33) | -1.16 |
| PF, median (IQR) | 85.00 (66.67 – 88.33) | 86.67 (80.00 – 93.33) | 7.03 |
| RF, median (IQR) | 100.00 (83.33 – 100.00) | 100.00 (83.33 – 100.00) | -7.36 |
| EF, median (IQR) | 91.67 (70.83 – 100.00) | 95.83 (83.33 – 100.00) | 7.68 |
| FA, median (IQR) | 22.22 (12.50 – 33.33) | 22.22 (11.11 – 33.33) | -1.74 |
| PA, median (IQR) | 16.67 (0.00 – 33.33) | 0.00 (0.00 – 33.33) | 1.39 |
| EORTC QLQ-PR25 | | | -2.85 |
| PRSAC, median (IQR) | 100.00 (83.33 – 100.00) | 100.99 (83.33 - 100.00) | -13.89 |
| PRSFU, median (IQR) | 66.67 (45.83 – 75.00) | 58.33 (37.50 – 79.17) | -1.16 |
| PRURI, median (IQR) | 20.83 12.50 – 30.21) | 20.83 (9.37 – 32.29) | -7.71 |
| PRBOW, median (IQR) | 8.33 (0.00 – 16.67) | 8.33 (0.00 – 8.33) | 6.08 |
| PRHT, median (IQR) | 22.22 (15.28 – 29.17) | 22.22 (11.11 – 38.89) | 0.00 |
| PRAID, median (IQR) | 0.00 (0.00 – 0.00) | 0.00 (00.00 – 00.00) | 0.00 |
| Cholesterol, median (IQR) | 4.40 (3.60 – 5.30) | 4.50 (3.75 – 5.40) | 1.43 |
| Triglyserides, median (IQR) | 1.27 (0.85 – 1.80) | 1.22 (0.93 – 1.71) | 6.45 |
| HDL, median (IQR) | 1.45 (1.21 – 1.66) | 1.54 (1.29 – 1.92) | 1.69 |
| LDL, median (IQR) | 2.50 (2.10 – 3.25) | 2.70 (2.10 – 3.75) | 1.56 |
| Glucose, median (IQR) | 6.20 (5.75 – 6.80) | 6.10 (5.75 – 7.05) | 0.88 |
| HbA1C, median (IQR) | 41.00 (36.50 – 44.50) | 39.00 (35.00 – 43.00) | -1.43 |

| | | | |
|---|---|---|---|
| Weight in kg, median (IQR) | 81.70 (72.55 – 100.25) | 83.20 (74.10 – 99.00) | 1.43 |
| BMI, median (IQR) | 25.90 (23.90 – 30.70) | 26.00 (24.60 – 30.20) | 6.45 |
| Whole body fat mass in kg, median (IQR) | 20.35 (14.65 – 29.58) | 21.10 (16.30 – 29.10) | 1.69 |
| Visceral fat mass in kg, median (IQR) | 14.50 (12.75 – 18.50) | 15.00 (14. 00 – 18.00) | 1.56 |
| Whole body lean mass in kg, median (IQR) | 59.45 (54.48 – 65.93) | 58.20 (54.60 – 66.20) | 0.88 |

*ERTC QLQ-30: Quality of Life (QL), Physical Functioning (PF), Role Functioning (RF), Emotional Functioning (EF), Fatigue (FA), and Pain (PA). PR25: Sexual Activity (PRSAC) and Sexual Functioning (PRSFU), Urinary Symptoms (PRURI), Bowel Symptoms (PRBOW), Hormonal Treatment-Related Symptoms (PRHTR), and Incontinence Aid Use (PRAID).*
